# Streamlining Large-Scale Genomic Data Management: Insights from the UK Biobank Whole-Genome Sequencing Data

**DOI:** 10.1101/2025.01.27.25321225

**Authors:** Xihao Li, Andrew R. Wood, Yuxin Yuan, Manrui Zhang, Yushu Huang, Gareth Hawkes, Robin N. Beaumont, Michael N. Weedon, Wenyuan Li, Xiaoyu Li, Xihong Lin, Zilin Li

**Affiliations:** Department of Biostatistics, University of North Carolina at Chapel Hill, Chapel Hill, NC, USA; Department of Genetics, University of North Carolina at Chapel Hill, Chapel Hill, NC, USA; Department of Biomedical and Clinical Sciences, Faculty of Health and Life Sciences, University of Exeter, Exeter, UK; School of Mathematics and Statistics and KLAS, Northeast Normal University, Changchun, Jilin, China; Department of Sociology, Tsinghua University, Beijing, China; Department of Big Data in Health Science, School of Public Health and Center of Clinical Big Data and Analytics of The Second Affiliated Hospital, Zhejiang University School of Medicine, Hangzhou, Zhejiang, China; Department of Biostatistics, Harvard T.H. Chan School of Public Health, Boston, MA, USA; Department of Statistics, Harvard University, Cambridge, MA, USA

## Abstract

Biobank-scale Whole-Genome Sequencing (WGS) studies are increasingly pivotal in unraveling the genetic bases of diverse health outcomes. However, managing and analyzing these datasets’ sheer volume and complexity presents significant challenges. We propose *vcf2agds*, an all-in-one toolkit that efficiently converts WGS data from Variant Call Format (VCF) format to the annotated Genomic Data Structure (aGDS) format, significantly reducing data size while supporting seamless genomic and functional data integration for comprehensive genetic analyses. The toolkit was applied to the UK Biobank 500k WGS data, resulting in twenty-three aGDS files, one for each chromosome, which collectively compressed 1,473.85 Tebibytes of pVCF data into 1.10 Tebibytes. Utilizing these aGDS files, we conducted a functionally informed rare variant association analysis of total cholesterol employing the *STAARpipeline* and detected 480 genome-wide significant coding and noncoding associations. Overall, *vcf2agds* offers a streamlined approach facilitating the efficient management and analysis of biobank-scale WGS data across hundreds of thousands of samples.

---

Over the past decade, the rapid advancements in whole-genome sequencing (WGS) technologies have generated vast amounts of data, enabling comprehensive assessment of associations between complex traits and both common and rare variants across coding and noncoding regions of the genome. Notably, the National Heart, Lung, and Blood Institute (NHLBI) Trans-Omics for Precision Medicine Program (TOPMed)^1^, the National Institutes of Health (NIH) All of Us Research Program^2^, and the UK Biobank^3^ have sequenced the whole genomes of approximately 200,000, 245,000, and 500,000 individuals, respectively. Together, these initiatives have identified over 1 billion genetic variants, presenting unprecedented opportunities and challenges for data management and analysis at the biobank scale. A widely used format for storing human genetic variation data is the Variant Call Format (VCF)^4^, which contains detailed information and genotype counts for all genomic variants across study participants. For instance, the UK Biobank released the 500k WGS jointly called variant data produced using GraphTyper (Data Field: 23374) in 151,561 blocks of project VCF (pVCF) files across the genome (**Supplementary Table 1**)^5^, with a total of 1473.85 Tebibyte (TiB) and an average of 9.96 Gibibytes (GiB) per file. However, such large size and inherent complexity of VCF files often hinder efficient downstream analyses, particularly in biobank-scale studies such as those involving the UK Biobank, which remains the only data format available since its release in November 2023.

To address the limitations of VCF files in large-scale genomic studies, several alternative data formats have been developed, including PLINK BED^6^, BGEN^7^, and Genomic Data Structure (GDS)^8^ format. Among these, the SeqArray GDS^9^ format is designed as a storage-efficient high-performance data format for WGS variant calls, utilizing a hierarchical structure to support efficient data access and seamless integration with several analytical pipelines. The annotated Genomic Data Structure (aGDS)^10^ format further extends the capabilities of SeqArray GDS by incorporating multi-faceted variant annotations to facilitate functionally-informed downstream analysis within an all-in-one file^11^.

Here we present *vcf2agds*, a streamlined and reproducible toolkit for processing the WGS data from VCF to aGDS files, with a case study using the UK Biobank WGS data (**Fig. 1**). The toolkit provides a four-step workflow, including VCF trimming, VCF merging, format conversion, and variant annotation. The first step (*vcf_trimmer*) involves reducing the complexity of VCF files using bcftools^12^ by removing potentially redundant fields and applying quality filters (e.g. excluding low quality variants with AAscore less than 0.5 assigned by GraphTyper^13^), that will not be used by downstream analyses. This preprocessing step minimizes file sizes and improves compatibility for merging, and users can customize this step by selecting specific fields and thresholds to retain the most relevant information for analysis. After trimming, the second step (*vcf_merger*) is to merge multiple VCF files into a unified dataset for each chromosome. Efficient merging ensures that the subsequent conversion to other file formats is streamlined and manageable, which is particularly critical for large-scale projects like the UK Biobank.

**Fig. 1:**
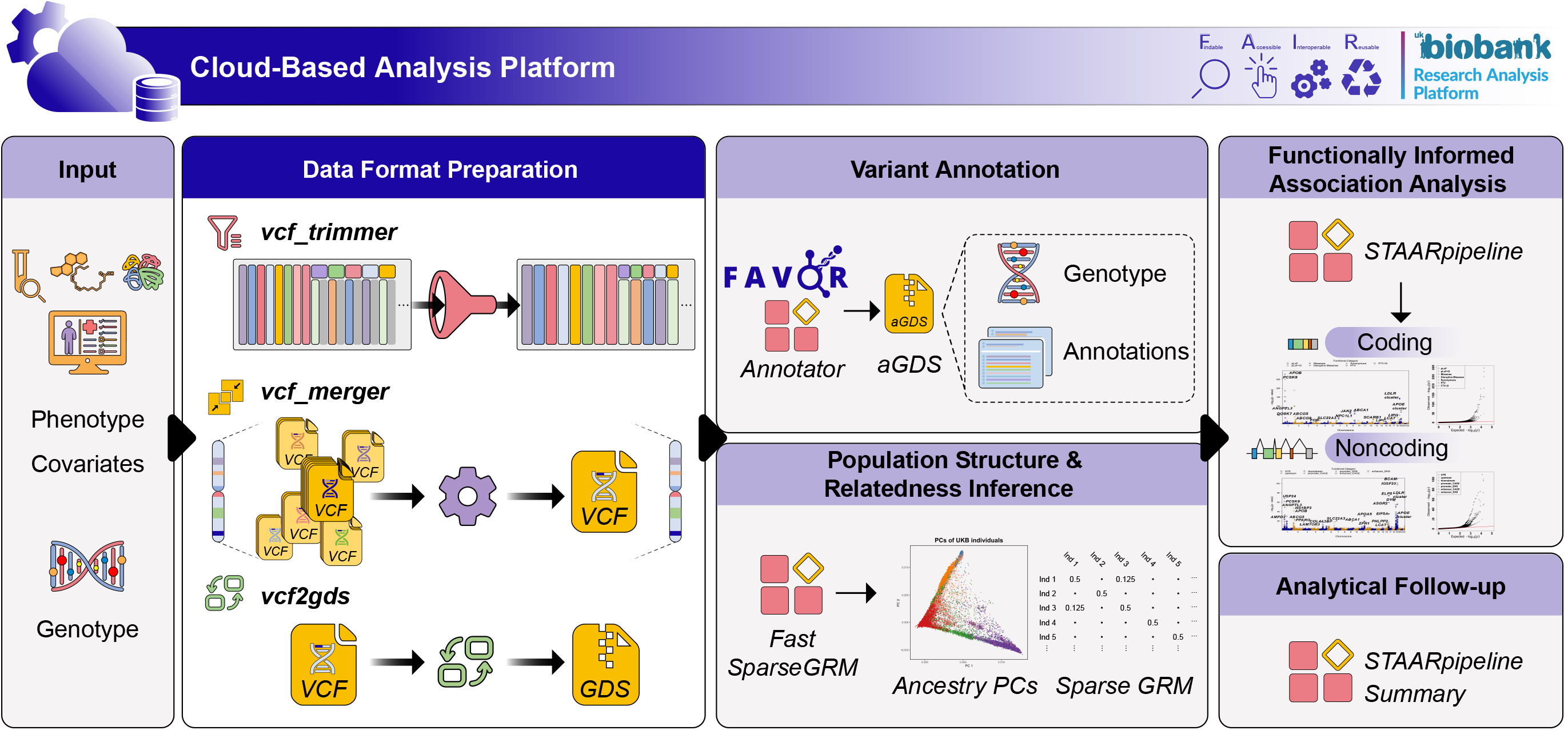
Overview of the cloud-based WGS analysis workflow. (i) prepare the input data, including genotypes, phenotypes and covariates; (ii) process WGS genotype data from VCF format to GDS format, using *vcf_trimmer, vcf_merger* and *vcf2gds* tools; (iii) functionally annotate all variants in the genome and generate aGDS files using *favorannotator*; (iv) calculate ancestry PCs and sparse GRM using *FastSparseGRM*; (v) perform functionally informed association analysis using *STAARpipeline*; (vi) provide result summarization, visualization and analytical follow-up using *STAARpipelineSummary*.

The third step (*vcf2gds*) converts the merged VCF files into GDS files using SeqArray^9^, which allows for efficient access to specific sample and/or variant subsets without loading the entire dataset into memory. The last step (*favorannotator*) performs functional annotation of the SeqArray GDS files into aGDS files through the Functional Annotation of Variants Online Resource (FAVOR) database^10^. The resulting aGDS files are equipped with both genotypes and variant annotations in the same file, providing detailed biological insights, including predicted functional impact, regulatory elements, protein function and conservation scores.

We applied *vcf2agds* to the 500k release of the UK Biobank WGS dataset, converting pVCF files (Data Field: 23374) to aGDS files (**Supplementary Table 1**). The original pVCF files, provided by the UK Biobank in 151,561 blocks, totaled 1,473.85 TiB. The aGDS files were generated as one file per chromosome, achieving a total size of only 1.10 TiB, representing a 1336x reduction compared to the pVCF files (**Supplementary Table 1**). As a downstream application of the utility of aGDS files, we conducted a comprehensive analysis of total cholesterol (TC) using variants that passed quality control in 500k WGS data (**Supplementary Table 2**). Leveraging the STAARpipeline for variant set analysis across diverse coding and noncoding genomic units^14^ (**Supplementary Note**), we identified 480 genome-wide significant associations. These associations were determined using Bonferroni-corrected significance thresholds of *α* = 0.05/(20,000 × 7) = 3.57 × 10^−7^ accounting for 7 different coding or noncoding masks across protein-coding genes, and *α* = 0.05/20,000 = 2.50 × 10^−6^ accounting for noncoding RNA genes. Among these, 200 associations were uncovered through gene-centric coding analyses, while 280 were identified through gene-centric noncoding analyses (**Supplementary Fig. 1, Supplementary Tables 3-4**). Further conditional analysis can be performed to detect putatively novel associations adjusting for the previously reported TC-associated variants and independent associations via aggregate conditioning^14,15^.

We further benchmarked the storage requirements of aGDS files using the 200k release of the UK Biobank WGS dataset, compared with pVCF files (Data Field: 24304), PLINK BED files (Data Field: 24305) and BGEN files (Data Field: 24306). The original pVCF files, PLINK BED and BGEN files totaled 533.62 TiB, 26.86 TiB and 0.87 TiB in size, respectively. The aGDS files, generated using *vcf2agds*, had a total size of 0.49 TiB, representing a 1084x reduction in size compared to the original pVCF files, and a compression ratio of 54.61x relative to PLINK BED files and 1.77x relative to BGEN files. Notably, the aGDS format uniquely integrates variant functional annotation data within the same file, making it the most compact and information-rich data structure among the formats compared (**Supplementary Table 5**). Although these benchmarks were conducted on data generated from joint variant calling using GraphTyper for both the 200k and 500k releases of the UK Biobank WGS data^5^, the proposed *vcf2agds* toolkit can seamlessly be applied to WGS data generated from joint variant calling with Illumina DRAGEN (Data Field: 24310) for the 500k release as well. These results underscore the flexibility and efficiency of the *vcf2agds* toolkit in managing large-scale genomic datasets, which only required to be performed once for all downstream analyses.

Overall, *vcf2agds* provides a streamlined solution for converting VCF files to aGDS format, enabling efficient management and analysis of biobank-scale WGS data of hundreds of thousand samples. As WGS datasets continue to expand in size and complexity, we hope that our proposed tools will provide an increasingly important role in unlocking the full potential of genomic research.

## Data Availability

Access to the UK Biobank resource is available via application (https://www.ukbiobank.ac.uk/). Development of software was undertaken under UK Biobank applications 9072 and 103356. Benchmarking results for UK Biobank WGS data were generated using the UK Biobank resource under applications 91486 and 52008. All analyses were conducted in the UK Biobank Research Analysis Platform (RAP, https://ukbiobank.dnanexus.com/).

## Code and data availability

*vcf2agds* is a suite of open-source software freely available at https://github.com/drarwood/vcf2agds_overview. Access to the UK Biobank resource is available via application (https://www.ukbiobank.ac.uk/). Development of software was undertaken under UK Biobank applications 9072 and 103356. Benchmarking results for UK Biobank WGS data were generated using the UK Biobank resource under applications 91486 and 52008. All analyses were conducted in the UK Biobank Research Analysis Platform (RAP, https://ukbiobank.dnanexus.com/), and the generated aGDS files will be made available as a ‘Returned Dataset’. STAARpipeline is implemented as an open-source R package available at https://github.com/xihaoli/STAARpipeline, and as an applet in UK Biobank RAP available at https://github.com/xihaoli/staarpipeline-rap.

## Acknowledgements

We would like to acknowledge Timothy M. Frayling at the University of Geneva for the contribution of funds for software development and testing on the DNAnexus Research Analysis Platform. This study is supported by the National Institute for Health and Care Research (NIHR) Exeter Biomedical Research Centre (BRC). The views expressed are those of the author(s) and not necessarily those of the NIHR or the Department of Health and Social Care. M.N.W. and R.N.B. are supported by MRC grant MR/Y003748/1. X. Lin is supported by the NIH grants R35-CA197449, R01-HL163560, U19-CA203654, U01-HG012064, and U01-HG009088.

## References

1. Taliun, D. et al. Sequencing of 53,831 diverse genomes from the NHLBI TOPMed Program. Nature 590, 290–299 (2021).

2. Bick, A.G. et al. Genomic data in the All of Us Research Program. Nature 627, 340–346 (2024).

3. Li, S., Carss, K.J., Halldorsson, B.V. & Cortes, A. Whole-genome sequencing of half-a-million UK Biobank participants. medRxiv, 2023.12.06.23299426 (2023).

4. Danecek, P. et al. The variant call format and VCFtools. Bioinformatics 27, 2156–2158 (2011).

5. Eggertsson, H.P. et al. Graphtyper enables population-scale genotyping using pangenome graphs. Nature Genetics 49, 1654–1660 (2017).

6. Purcell, S. et al. PLINK: A Tool Set for Whole-Genome Association and Population-Based Linkage Analyses. The American Journal of Human Genetics 81, 559–575 (2007).

7. Band, G. & Marchini, J. BGEN: a binary file format for imputed genotype and haplotype data. bioRxiv, 308296 (2018).

8. Zheng, X. et al. A high-performance computing toolset for relatedness and principal component analysis of SNP data. Bioinformatics 28, 3326–3328 (2012).

9. Zheng, X. et al. SeqArray—a storage-efficient high-performance data format for WGS variant calls. Bioinformatics 33, 2251–2257 (2017).

10. Zhou, H. et al. FAVOR: functional annotation of variants online resource and annotator for variation across the human genome. Nucleic Acids Research 51, D1300–D1311 (2023).

11. Li, X. et al. Dynamic incorporation of multiple in silico functional annotations empowers rare variant association analysis of large whole-genome sequencing studies at scale. Nature Genetics 52, 969–983 (2020).

12. Li, H. A statistical framework for SNP calling, mutation discovery, association mapping and population genetical parameter estimation from sequencing data. Bioinformatics 27, 2987–2993 (2011).

13. Halldorsson, B.V. et al. The sequences of 150,119 genomes in the UK Biobank. Nature 607, 732–740 (2022).

14. Li, Z. et al. A framework for detecting noncoding rare-variant associations of large-scale whole-genome sequencing studies. Nature Methods 19, 1599–1611 (2022).

15. Hawkes, G. et al. Whole-genome sequencing in 333,100 individuals reveals rare non-coding single variant and aggregate associations with height. Nature Communications 15, 8549 (2024).

