## Supplementary Information for "Streamlining Large-Scale Genomic Data Management: Insights from the UK Biobank Whole-Genome Sequencing Data"

**Supplementary Note**

**Analysis of total cholesterol (TC) phenotype in the UK Biobank 500k WGS data**

We used the generated aGDS files for WGS data of 490,549 UK Biobank participants from the pVCF files (Data Field: 23374) and followed the same quality control procedure in the previous study of UK Biobank WGS data^1^. We kept all variants indicated by the FILTER label and AAscore greater than 0.5, where AAscore was generated by GraphTyper^2^, the software used by the UK Biobank to perform genotype calling. Variants were further filtered by excluding those that failed the Hardy-Weinberg equilibrium test ($P<1\times{10}^{-100}$) or had a call rate below 90%^3,4^. We harmonized the total cholesterol trait (Data Field: 30690), where TC was adjusted by dividing the value by 0.8 among individuals reporting lipid lowering medication use or statin use (Data Field: 20003). A total of 462,458 individuals had data on TC.

We fit a linear regression model adjusting for age, age^2^, sex, and the first ten ancestral principal components. Residuals were then rank-based inverse-normal transformed and multiplied by the standard deviation. We next fit a linear mixed model (LMM) for the rank normalized residuals of TC, adjusting for age, age^2^, sex, and the ten ancestral principal components, and a variance component for an empirically-derived sparse kinship matrix using FastSparseGRM^5^ to account for population structure and relatedness.

We next applied STAARpipeline^6^ to perform variant set analyses for rare variants (MAF < 1%), including gene-centric analysis of protein-coding genes using seven coding variant functional categories; seven noncoding variant functional categories and rare variants in noncoding RNA (ncRNA) genes. Our analysis was performed on the UK Biobank Research Analysis Platform (RAP).

In the gene-centric coding analysis, 200 genome-wide significant associations were identified at the Bonferroni-corrected level $0.05/\left( 20,000\times7 \right)=3.57\times{10}^{-7}$ (**Supplementary Fig.1 a-b and Supplementary Table 3**). These associations included well-known genes such as *PCSK9*, *APOB*, the *LDLR* cluster, and the *APOE* cluster. For the gene-centric noncoding analysis of protein-coding genes, 271 genome-wide significant associations were detected at the Bonferroni-corrected level $0.05/\left( 20,000\times7 \right)=3.57\times{10}^{-7}$ (**Supplementary Fig.1 c-d and Supplementary Table 4**). Among these, notable associations involved genes such as *USP24*, *PCSK9*, *APOB*, the *LDLR* cluster, and the *APOE* cluster. In the gene-centric noncoding analysis of ncRNA genes, we identified 9 significant associations at the genome-wide significance level $0.05/20,000=2.50\times{10}^{-6}$ (**Supplementary Fig.1 e-f and Supplementary Table 4**).

**References**

1.	Halldorsson, B.V. *et al.* The sequences of 150,119 genomes in the UK Biobank. *Nature* **607**, 732-740 (2022).

2.	Eggertsson, H.P. *et al.* Graphtyper enables population-scale genotyping using pangenome graphs. *Nature Genetics* **49**, 1654-1660 (2017).

3.	Zhao, Y. *et al.* Population scale whole genome sequencing provides novel insights into cardiometabolic health. *medRxiv*, 2024.05.27.24307970 (2024).

4.	Li, S., Carss, K.J., Halldorsson, B.V. & Cortes, A. Whole-genome sequencing of half-a-million UK Biobank participants. *medRxiv*, 2023.12.06.23299426 (2023).

5.	Lin, X., Dey, R., Li, X. & Li, Z. Scalable analysis of large multi-ancestry biobanks by leveraging sparse ancestry-adjusted sample-relatedness. *Res Sq* (2024).

6.	Li, Z. *et al.* A framework for detecting noncoding rare-variant associations of large-scale whole-genome sequencing studies. *Nature Methods* **19**, 1599-1611 (2022).

**Supplementary Figures**

**Supplementary Figure 1. Manhattan plots and Q-Q plots for gene-centric coding analysis and noncoding analysis of TC. (a)** Manhattan plots for gene-centric coding analysis. The horizontal line indicates a genome-wide STAAR-O *P*-value threshold of $3.57\times{10}^{-7}$. The significant threshold is defined by multiple comparisons using the Bonferroni correction ($0.05/\left( 20,000\times7 \right)=3.57\times{10}^{-7}$). **(b)** Quantile-quantile plots for gene-centric coding analysis. **(c)** Manhattan plots for gene-centric noncoding analysis. The horizontal line indicates a genome-wide STAAR-O *P*-value threshold of $3.57\times{10}^{-7}$. The significant threshold is defined by multiple comparisons using the Bonferroni correction ($0.05/\left( 20,000\times7 \right)=3.57\times{10}^{-7}$). **(d)** Quantile-quantile plots for gene-centric noncoding analysis. **(e)** Manhattan plots for ncRNA analysis. The horizontal line indicates a genome-wide STAAR-O *P*-value threshold of $2.50\times{10}^{-6}$. The significant threshold is defined by multiple comparisons using the Bonferroni correction ($0.05/20,000=2.50\times{10}^{-6}$). **(f)** Quantile-quantile plots for ncRNA analysis. In panels, **(a), (c)** and **(e)**, the chromosome number are indicated by the colors of dots. In all panels, STAAR-O is a two-sided test.

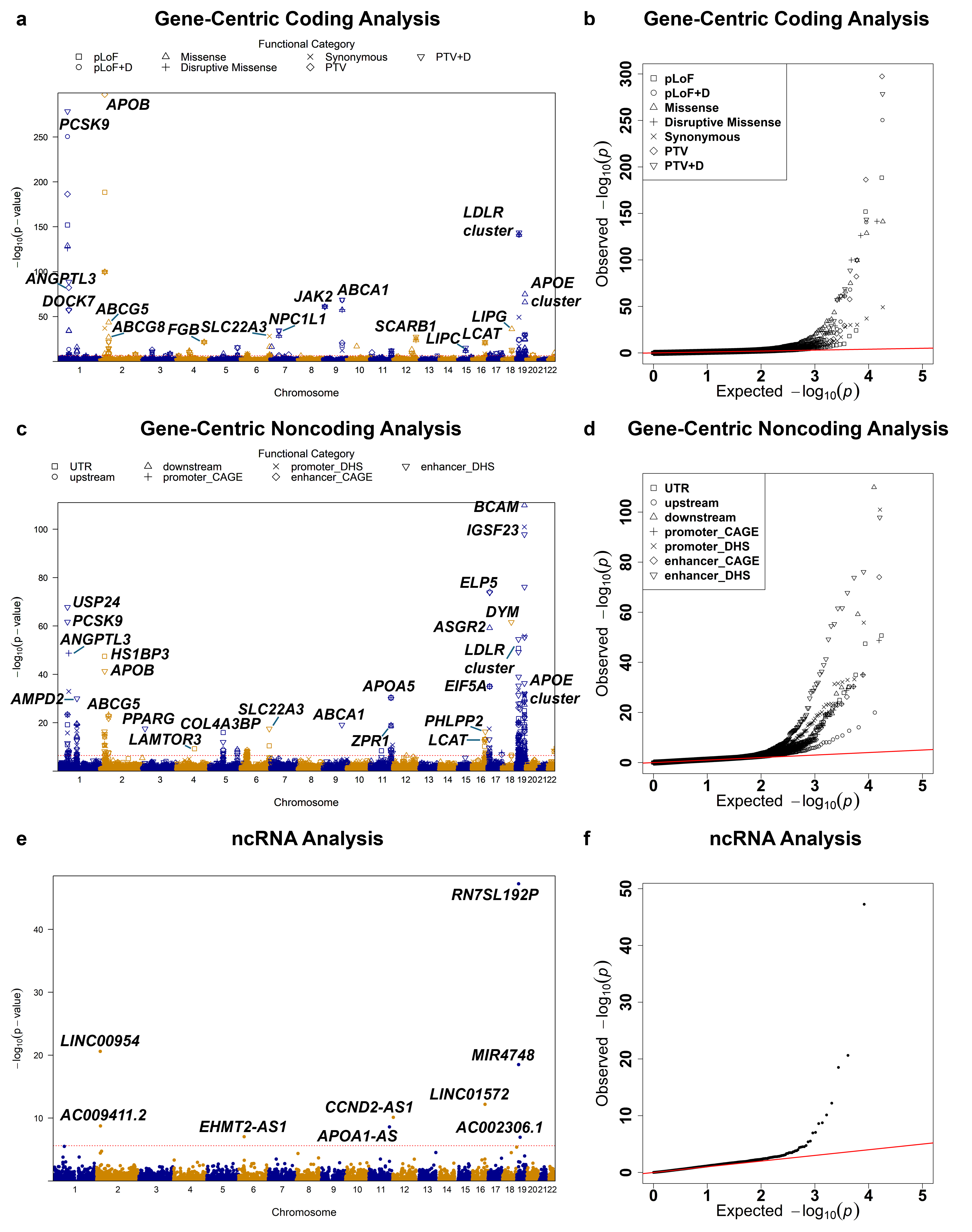
